# Trends in congenital clubfoot prevalence and co-occurring anomalies during 1994-2021 in Denmark: A nationwide register-based study of 1,315,282 live born infants

**DOI:** 10.1101/2023.05.11.23289837

**Authors:** Paula L. Hedley, Ulrik Lausten-Thomsen, Kristin M. Conway, Klaus Hindsø, Paul A. Romitti, Michael Christiansen

**Author notes:** **Corresponding author:** Paula L. Hedley, PhD, MPH, Department for Congenital Disorders, Statens Serum Institut, 5 Artillerivej DK2300S, Copenhagen, Denmark.

## Abstract

**Background:** Congenital talipes equinovarus (clubfoot) is a common musculoskeletal anomaly, with a suspected multifactorial etiopathogenesis. Herein, we used publicly available data to ascertain liveborn infants with clubfoot delivered in Denmark during 1994–2021, and to classify co-occurring congenital anomalies, estimate annual prevalence, and compare clubfoot occurrence with maternal smoking rates, a commonly reported risk factor. Characterizing this nationwide, liveborn cohort provides a population-based resource for etiopathogenic investigations and life course surveillance.

**Methods:** This case-cohort study used data from the Danish National Patient Register and Danish Civil Registration System, accessed through the publicly available Danish Biobank Register, to identify 1,315,282 liveborn infants delivered during 1994–2021 in Denmark to Danish parents. Among these, 2,358 infants (65.1% male) were ascertained with clubfoot and classified as syndromic (co-occurring chromosomal, genetic, or teratogenic syndromes) and nonsyndromic (isolated or co-occurring multiple congenital anomalies [MCA]). Annual prevalence estimates and corresponding 95% confidence intervals (CIs) for children with nonsyndromic clubfoot were estimated using Poisson regression and compared with population-based, maternal annual smoking rates obtained from publicly available resources.

**Results:** Infants most often presented with nonsyndromic clubfoot (isolated=84.6%; MCA=10.9%); limb and heart anomalies were the most frequently identified MCAs. Prevalence (per 1,000 liveborn infants) was 1.52 (CI 1.45 – 1.58) for isolated and 0.19 (CI 0.17 – 0.22) for MCA clubfoot. Prevalence estimates for both isolated and MCA clubfoot remained relatively stable during the study period, despite marked decreases in population-based maternal smoking rates.

**Conclusions:** From 1994-2021, prevalence of nonsyndromic clubfoot in Denmark was relatively stable. Reduction in population-level maternal smoking rates did not seem to impact prevalence estimates, providing some support for the suspected multifactorial etiopathogenesis of this anomaly. This nationwide, liveborn cohort, ascertained and clinically characterized using publicly available data from the Danish Biobank Register, provides a population-based clinical and biological resource for future etiopathogenic investigations and life course surveillance.

## Background

Congenital talipes equinovarus (commonly referred to as clubfoot) is the most common, major musculoskeletal anomaly affecting newborns, with an estimated occurrence of 1-2 per 1,000 births [1]. This condition is more frequently observed in males and bilateral presentation is slightly more frequent than unilateral [1]. The most common treatment for clubfoot is the Ponseti method, which involves serial stretching and casting [2]; often followed by tenotomy [3]. Untreated clubfoot results in serious disability [3].

Approximately 80% of individuals with clubfoot present as isolated; the remainder present with other congenital anomalies, including syndromic (chromosomal, genetic, or teratogenic) or nonsyndromic (multiple congenital anomalies [MCAs]) phenotypes [4]. Clubfoot presenting with neuromuscular anomalies (e.g., neural tube defects, arthrogryposis), bilateral renal agenesis, and Potter sequence are considered secondary to these anomalies [4].

Clubfoot etiopathogenesis is poorly understood, but likely caused by a combination of gene variants and environmental (broadly defined) exposures [5]. Excess of clubfoot among males and certain racial/ethnic groups [6], together with the proportion of bilateral diagnoses, suggest strong genetic contributions. Additionally, evidence of a genetic component among individuals with isolated clubfoot is suggested by the heritability of 30% estimated in a Danish twin study [5]. Several environmental exposures have also been associated with clubfoot, most consistently maternal cigarette smoking [7]. Furthermore, even with correction in infancy, the varying propensity of relapse observed among children provides additional evidence that clubfoot has a heterogenous etiopathogenesis [8].

In this study, we utilize publicly available data from nationwide registers in Denmark to investigate the prevalence of clubfoot diagnoses among live births during 1994-2021. We describe congenital anomalies co-occurring with clubfoot and estimate annual prevalence during this period as well as during a restricted period (2010-2021), following establishment of clubfoot specialist centers [9]. Additionally, we compare prevalence estimates with population-based, maternal annual smoking rates as well as with the European network of congenital anomalies registers (EUROCAT) Danish sub-population from the Region of Southern Denmark. Through this effort, we aim to establish a nationwide, liveborn cohort for future etiopathogenic investigations and life course surveillance of clinical outcomes.

## Methods

We aim to ascertain and clinically characterize a national cohort of Danish clubfoot cases, using publicly available nationwide register data.

The Danish Biobank Register contains information from the Danish Civil Registration System (date of birth, country of birth, and country of birth of both parents) and the Danish National Patient Register (diagnostic codes and dates of diagnosis) for all individuals with specimens stored in the Danish National Biobank. Using the publicly available Danish Biobank Register online interface [10], we ascertained the number of infants, born during 1^st^ January 1994 through 31^st^ December 2021 and diagnosed with congenital clubfoot (ICD-10-DK: DQ660 – talipes equinovarus or DQ663B – clubfoot, unspecified) within one year of birth using data from individuals sampled for neonatal screening. Although terminations of pregnancy, fetal deaths, and very early neonatal deaths are naturally excluded, the coverage for neonatal screening in Denmark is close to 100% [11]; consequently, our study sample represents a nationwide cohort of all infants alive at time of screening (days 5-7 in 1994-2008, and days 2-3 in 2009-2021).

To reduce etiopathogenic heterogeneity, case children diagnosed with neural tube defects (ICD-10-DK: DQ00, DQ01, DQ05), bilateral renal agenesis (ICD-10-DK: DQ60.1), Potter sequence (ICD-10-DK: DQ60.6), or arthrogryposis multiplex congenita (ICD-10-DK: DQ74.3) were excluded, because clubfoot secondary to these diagnoses would be considered etiopathogenically different from primary (idiopathic) clubfoot [4]. The cohort was also limited to children born in Denmark to Danish parents (both parents born in Denmark). For the sake of comparison, the prevalence of primary clubfoot in children born to parents where neither, one, or both parents were themselves born in Denmark is shown in Supplementary Figure 1.

## Statistics

Eligible, congenital clubfoot case children were classified as isolated (no additional, major congenital anomalies) or presenting with co-occurrence of major congenital anomalies, using a hierarchical grouping of chromosomal anomalies, genetic syndromes, teratogenic syndromes, and nonsyndromic, major anomalies (ICD-10-DK codes shown in Supplementary Table 1). Frequencies and proportions for descriptive characteristics of each group were calculated. Subsequent analyses were restricted to children with nonsyndromic clubfoot (i.e., isolated clubfoot and clubfoot with MCAs).

A trend analysis was performed using Poisson regression for isolated and MCA clubfoot cases. Pearson correlation was performed to assess the relationship between numerical variables. Prevalence of clubfoot was estimated by dividing the total number of infants diagnosed with clubfoot by the total number of infants sampled for neonatal screening. Annual trends, since the establishment of clubfoot specialist centers in Denmark (2010-2021) [9], of the prevalence of clubfoot were examined using Poisson regression. Results from the Poisson regression model were used to estimate prevalence rate ratios (PRRs) and 95% confidence intervals (CIs).

To enable comparison with previous studies, annual counts of liveborn children with nonsyndromic clubfoot from the EUROCAT Danish sub-population (The Region of Southern Denmark) were extracted [12]. Furthermore, population data and data pertaining to municipal area in km^2^ were extracted from Statistics Denmark [13], from 2022, to assess the median (range) population density of municipalities in The Region of Southern Denmark and throughout Denmark.

With maternal smoking during pregnancy repeatedly reported as a risk factor for clubfoot, prevalence estimates were compared with population-based, annual maternal smoking rates from 1999-2021, which were extracted from the Medical Birth Register [14] using the publicly available online interface esundhed.dk [15].

All analyses were performed using R version 4.2.2.

## Results

During 1994-2021, 1,696,353 live-born infants were delivered in Denmark and sampled for neonatal screening, 1,315,282 of these were delivered to Danish parents (both parents were themselves born in Denmark). There were no diagnoses of anencephaly, encephalocele, bilateral renal agenesis, or Potter sequence identified among the case children. After excluding case children with spina bifida (n=32), arthrogryposis multiplex congenita (n=40), or with co-occurring spina bifida and arthrogryposis multiplex congenita (n=1), 2,358 (65.1% male) case children with primary clubfoot were ascertained. Phenotype classification of case children showed 1,995 (84.6%) infants with isolated clubfoot, 45 (1.9%) with a chromosomal anomaly, 57 (2.4%) with a genetic syndrome, five (0.2%) with a teratogenic syndrome, and 256 (10.9%) with at least one major congenital anomaly in another organ system (Table 1, Figure 1), the most frequent of which were limb (n=121; 47.3%) and cardiac (n=59; 23.0%) anomalies (Table 1). Restricting case children to those with nonsyndromic clubfoot (Figure 1), produced an overall prevalence (per 1,000 livebirths) of 1.71 (CI 1.64 – 1.78) (Table 1), with higher estimates for males (2.16, CI 2.05 – 2.27) than females (1.24, CI 1.15 – 1.33) (Figure 1). The annual prevalence estimates for nonsyndromic clubfoot, stratified by males and females are shown in Figure 2.

**Table 1:**
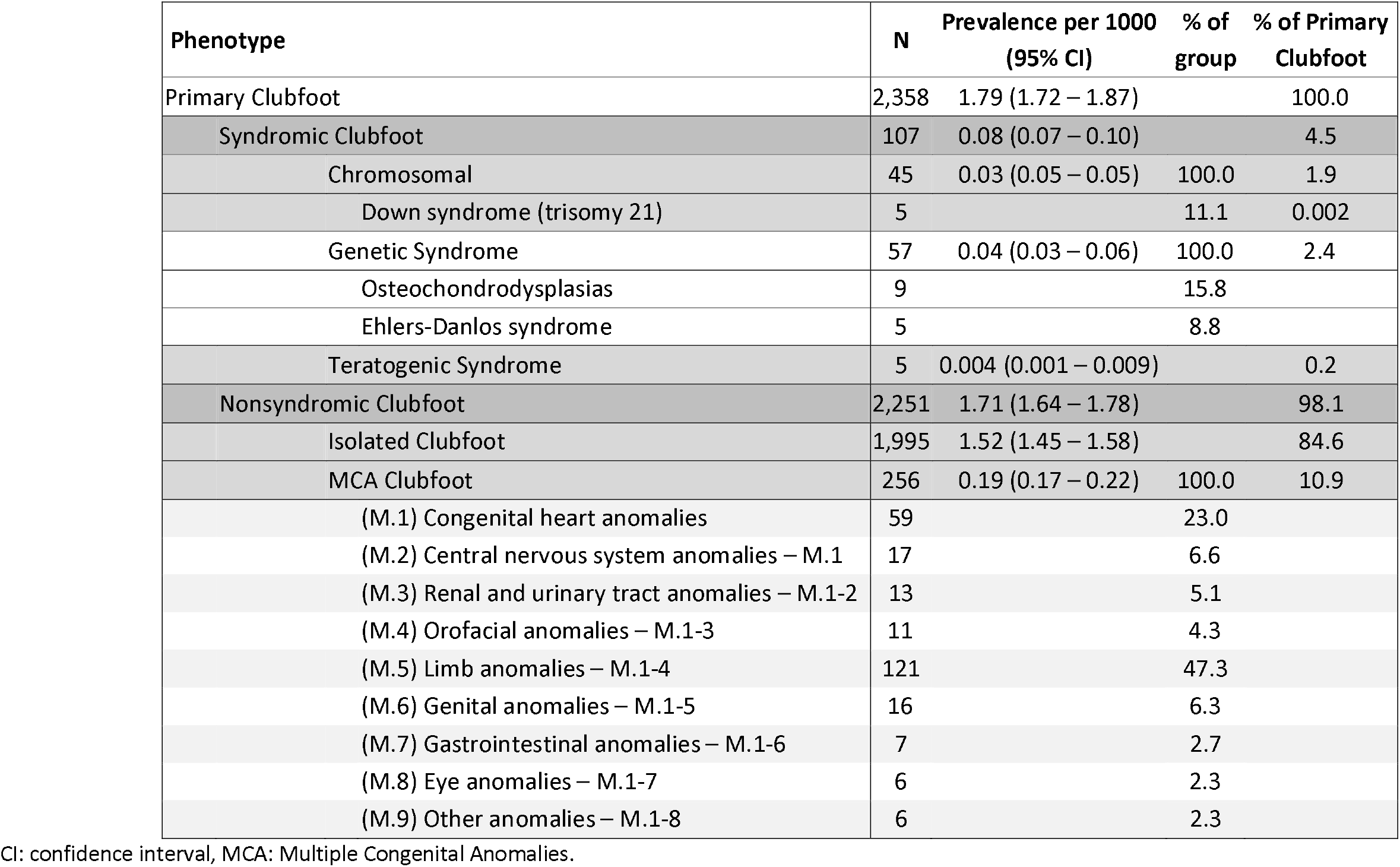
The number and prevalence per 1,000 infants with congenital clubfoot associated with syndromic and nonsyndromic clubfoot cases diagnosed in Denmark during 1994 – 2021. Diagnoses were assessed hierarchically in the order shown in Supplementary Table 1.

| Phenotype | N | Prevalence per 1000<br>(95% CI) | % of<br>group | % of Primary<br>Clubfoot |
| --- | --- | --- | --- | --- |
| Primary Clubfoot | 2,358 | 1.79 (1.72 – 1.87) |  | 100.0 |
| Syndromic Clubfoot | 107 | 0.08 (0.07 – 0.10) |  | 4.5 |
| Chromosomal | 45 | 0.03 (0.05 – 0.05) | 100.0 | 1.9 |
| Down syndrome (trisomy 21) | 5 |  | 11.1 | 0.002 |
| Genetic Syndrome | 57 | 0.04 (0.03 – 0.06) | 100.0 | 2.4 |
| Osteochondrodysplasias | 9 |  | 15.8 |  |
| Ehlers-Danlos syndrome | 5 |  | 8.8 |  |
| Teratogenic Syndrome | 5 | 0.004 (0.001 – 0.009) |  | 0.2 |
| Nonsyndromic Clubfoot | 2,251 | 1.71 (1.64 – 1.78) |  | 98.1 |
| Isolated Clubfoot | 1,995 | 1.52 (1.45 – 1.58) |  | 84.6 |
| MCA Clubfoot | 256 | 0.19 (0.17 – 0.22) | 100.0 | 10.9 |
| (M.1) Congenital heart anomalies | 59 |  | 23.0 |  |
| (M.2) Central nervous system anomalies – M.1 | 17 |  | 6.6 |  |
| (M.3) Renal and urinary tract anomalies – M.1-2 | 13 |  | 5.1 |  |
| (M.4) Orofacial anomalies – M.1-3 | 11 |  | 4.3 |  |
| (M.5) Limb anomalies – M.1-4 | 121 |  | 47.3 |  |
| (M.6) Genital anomalies – M.1-5 | 16 |  | 6.3 |  |
| (M.7) Gastrointestinal anomalies – M.1-6 | 7 |  | 2.7 |  |
| (M.8) Eye anomalies – M.1-7 | 6 |  | 2.3 |  |
| (M.9) Other anomalies – M.1-8 | 6 |  | 2.3 |  |

CI: confidence interval, MCA: Multiple Congenital Anomalies.

**Figure 1.**
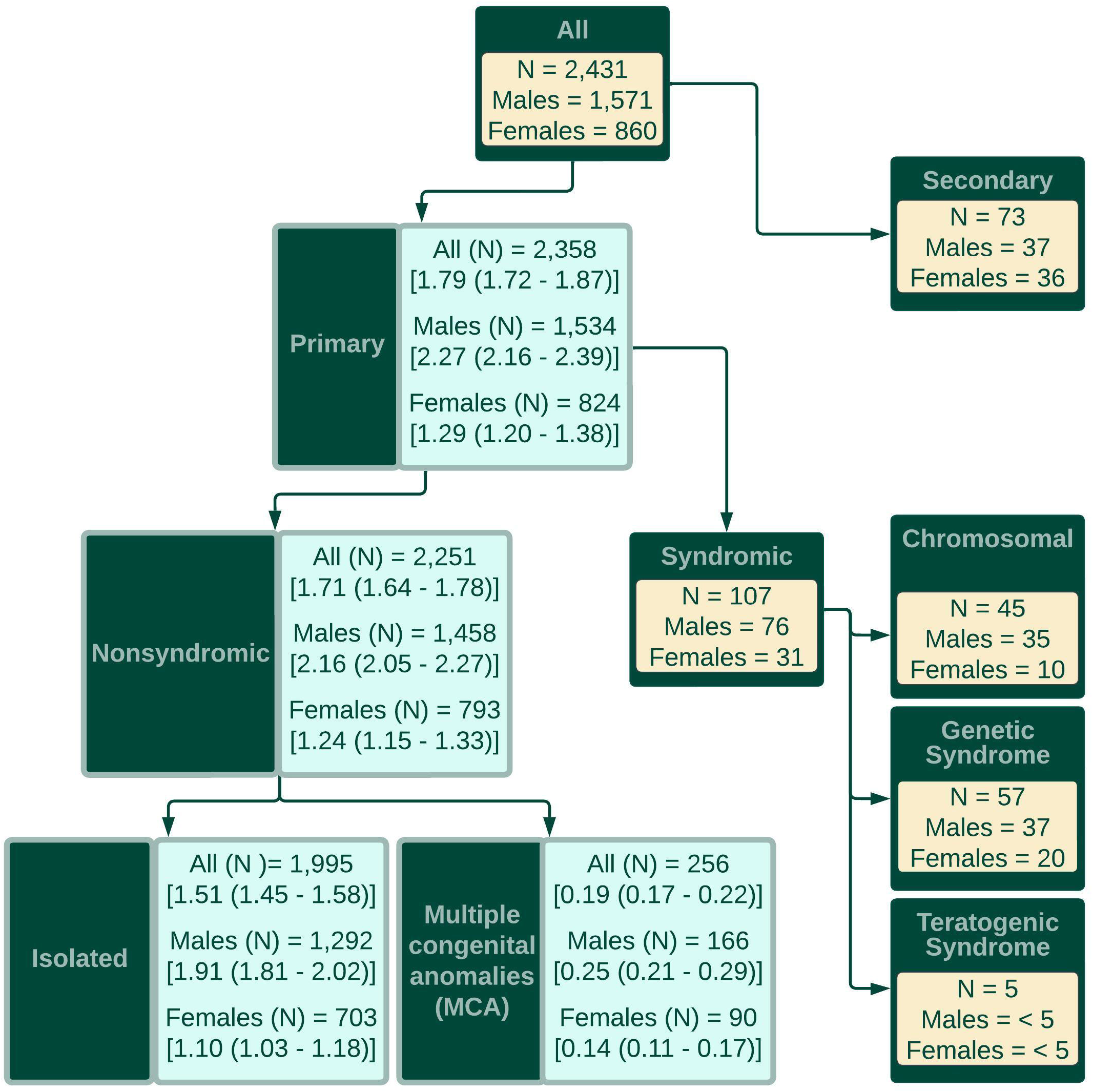

**Figure 2.**
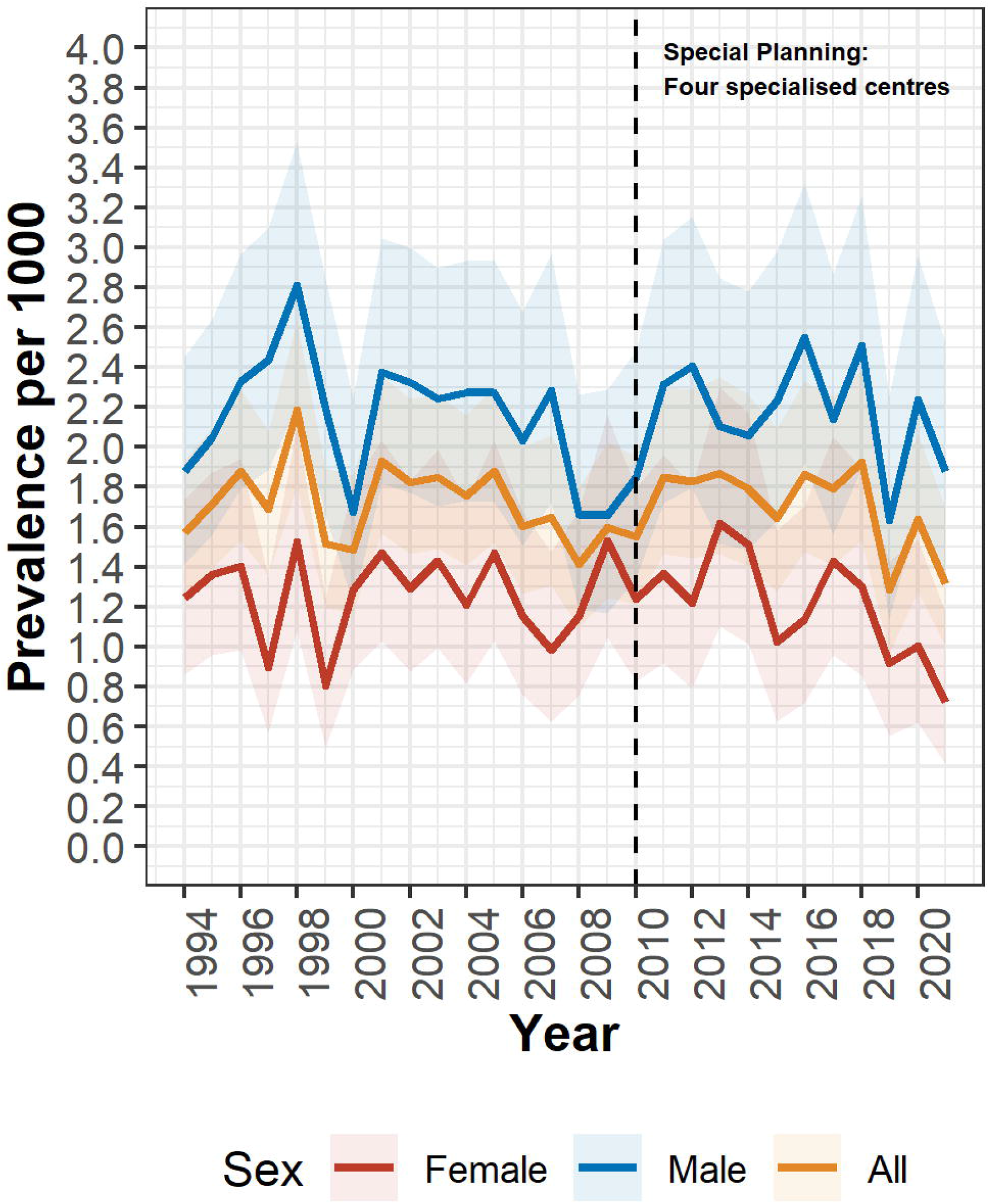

Trend analysis showed no statistically significant change in annual prevalence across the study period (data not shown). Similarly, restricting the analysis to 2010-2021 (the years following the implementation of specialized clubfoot treatment centers [9]) showed that the prevalence estimates for both isolated and MCA clubfoot were not significantly associated with the year of birth, indicating a relatively stable prevalence rate (Table 2).

**Table 2:** Crude and adjusted (for infant sex) prevalence rate ratios for isolated clubfoot and clubfoot with multiple congenital anomalies by year of birth and maternal smoking for the years following implementation of specialized clubfoot treatment centers in Denmark. 95% confidence interval ranges are presented in brackets and significance levels are indicated (* p<0.05, ** p<0.01, *** p<0.001).

**Isolated Clubfoot (2010-2021)**
| Coefficient | PRR<br>(95% CI) | PRR<br>(95% CI) | PRR<br>(95% CI) | aPRR<br>(95% CI) | aPRR<br>(95% CI) |
| --- | --- | --- | --- | --- | --- |
| Sex: Male | 1.76 ***<br>(1.52 – 2.04) |  |  | 1.76 ***<br>(1.52 – 2.04) | 1.75 ***<br>(1.52 – 2.03) |
| Year of birth |  | 0.99<br>(0.95 – 1.02) |  | 0.99<br>(0.97 – 1.00) |  |
| Maternal smoking rate |  |  | 1.00<br>(1.00 – 1.00) |  | 1.00<br>(1.00 – 1.00) |

**MCA Clubfoot (2010-2021)**
| Coefficient | PRR<br>(95% CI) | PRR<br>(95% CI) | PRR<br>(95% CI) | aPRR<br>(95% CI) | aPRR<br>(95% CI) |
| --- | --- | --- | --- | --- | --- |
| Sex: Male | 2.30 ***<br>(1.37 – 4.01) |  |  | 2.30 ***<br>(1.37 – 4.01) | 2.25 ***<br>(1.34 – 3.94) |
| Year of birth |  | 0.96<br>(0.88 – 1.04) |  | 0.96<br>(0.89 – 1.03) |  |
| Maternal smoking rate |  |  | 1.00<br>(1.00 – 1.00) |  | 1.00<br>(1.00 – 1.00) |

aPRR: adjusted prevalence rate ratio, CI: confidence interval, MCA: multiple congenital anomalies, PRR: prevalence rate ratio.

Comparison of nationwide prevalence estimates for liveborn children with nonsyndromic clubfoot with those from the Region of Southern Denmark included in EUROCAT showed that the region of Southern Denmark had a lower prevalence of nonsyndromic clubfoot than our nationwide estimate (1.14, CI 0.97 – 1.34 vs 1.71, CI 1.64 – 1.78, respectively). The annual prevalence estimates for nonsyndromic clubfoot for both the Region of Southern Denmark and Denmark as a whole are shown in Figure 3. Additionally, comparison between the population density of the municipal centers of Denmark showed that The Region of Southern Denmark had a lower median population density (persons/km^2^) than the municipal centers in Denmark as a whole (median 81, range 29 – 680 vs median 121, range 15 – 12,030, respectively).

**Figure 3.**
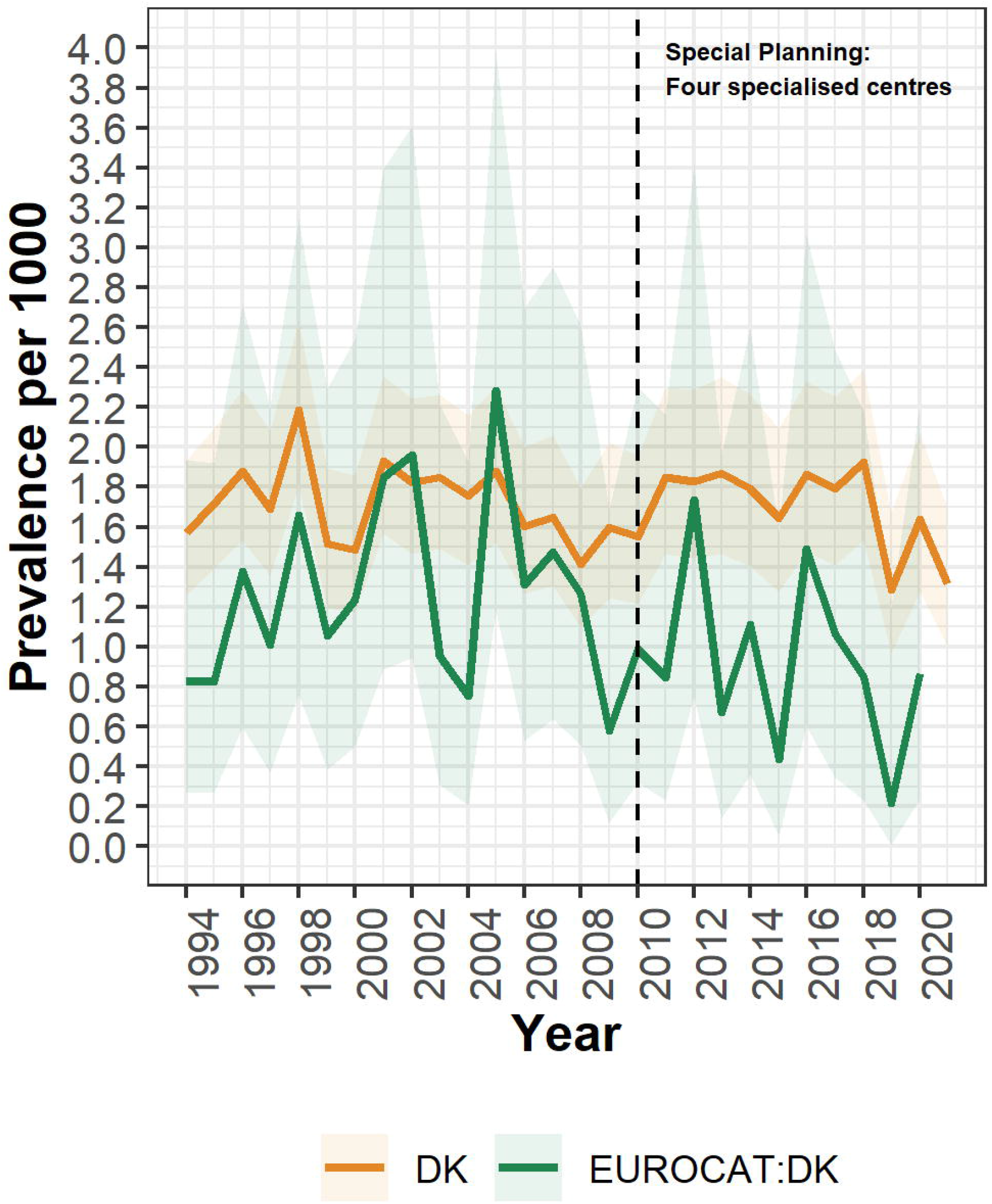

As anti-tobacco policies have been adopted and implemented in Denmark, maternal smoking has dropped continuously over the past 20 years (Supplementary Figure 2) with reported maternal smoking and year of birth being negatively correlated (r = -0.99 (CI -0.99 – -0.97)). As such, maternal smoking and year of birth were modelled separately. The rate of maternal smoking from 1999-2021 was not associated with either isolated or MCA clubfoot (Table 2).

## Discussion

We have ascertained and clinically characterized the most recently established nationwide clubfoot cohort in Denmark using exclusively publicly available data. For the birth period 1994-2021, we ascertained 2,358 liveborn infants with primary clubfoot of which 2,251 presented with nonsyndromic clubfoot. The overall prevalence (per 1,000 live births) for nonsyndromic clubfoot during this birth period was 1.71. This estimate is higher than the corresponding estimate of 1.14 reported to EUROCAT for the sub-population in The Region of Southern Denmark that covers 21% of the Danish population [12]. This discrepancy in prevalence may reflect true regional and/or ethnic differences, as our cohort was a nationwide sample limited to live births delivered in Denmark to Danish parents. Furthermore, as data on postural clubfoot was not available from the Danish Biobank Register, we did not specifically exclude these cases, which may have increased the number of misclassified cases in our cohort. As population density (persons/km^2^) was positively associated with clubfoot prevalence in a previous Danish study [16], we compared the population density of the municipal centers in The Region of Southern Denmark with the municipal centers in Denmark as a whole. The higher population density for Denmark as a whole may explain, in part, the differences in prevalence reported between our study and the Danish sub-population [12]. Our higher estimated prevalence cannot be explained by differences in delivery types included in each respective study. Because the infant would have had to survive to be sampled for neonatal screening, we did not have data on clubfoot among neonatal deaths that occurred within 24 hours of birth (2.4/1,000 deaths in Denmark during 1994-2021) [15]. These very early neonatal deaths would have been included as liveborn case children in the Danish subpopulation of EUROCAT [12]. Despite these methodologic differences, the comparison between our population and the EUROCAT Danish subpopulation indicates a reasonable overlap between annual prevalence estimates (Figure 3).

Fewer than one-half of the proportion of clubfoot cases in this study presented with a chromosomal anomaly when compared to a EUROCAT study [4] and a French study [17]. In particular, both previous studies saw a larger proportion of trisomy 18 and trisomy 13 cases. The differences in contributing chromosomal anomalies between studies probably reflects our inability to include clubfoot diagnoses among fetal losses (stillbirths [4.2/1,000 of all births in Denmark during 1994-2021] [15], as well as spontaneous abortions and terminations of pregnancy [0.5/1,000 of all births in Denmark during 1994-2021 occurring between gestational age 12 weeks 0 days to 21 weeks 6 days] [15]) and early neonatal deaths prior to sampling in this study. Also, case children with teratogenic syndrome cases were rare in our cohort precluding comparisons with other studies. Additionally, we excluded case children with clubfoot secondary to arthrogryposis, spina bifida, anencephaly, encephalocele, Potter sequence, and renal agenesis [4, 6], making comparisons to studies which did not exclude such cases [17] complicated. Furthermore, our cohort contained proportionally more case children with MCAs than those identified in the EUROCAT study [4]. We identified additional limb anomalies to be the most frequently co-occurring MCA, followed by congenital heart defects.

Multiple pathways are involved in the development of the lower limb. Consequently, several risk factors have been associated with clubfoot; male sex [4, 6], maternal smoking [18], and genetics [5] are the risk factors with the most robust evidence of association [19]. Maternal obesity [6, 7], amniocentesis or chorionic villus sampling [7, 20], population density [16], and SSRI exposure [7, 21], have also been associated with an increased risk of delivering an infant with clubfoot. We were able to evaluate sex and maternal smoking rates in our population and observed, as others have reported, that clubfoot occurs more frequently in males than females [4, 7, 22]. Maternal smoking rates have decreased considerably since 1999 (Supplementary Figure 2) and were significantly correlated with year of birth, but not with prevalence of congenital clubfoot. Other risk markers, for instance a reduction in SSRI use in pregnancy that has been reported between 2009 and 2016 [23], may also contribute to variation in clubfoot prevalence over time. However, it is worth noting that a Danish study assessing the risk of congenital anomalies among children born to women who redeemed their prescription for SSRIs during pregnancy reported no association with either a congenital anomaly of the limb generally [24], or clubfoot specifically [25].

A limitation of our study is that, by examining children alive and well at the time of sampling for neonatal screening, we are unable to report the prevalence of clubfoot among fetal and early neonatal deaths. However, the children ascertained in this study represent the population utilizing the Danish healthcare system (clubfoot treatment is not initiated before the child is old enough to be registered for neonatal screening), it is, therefore, important to characterize this population. Another limitation is that these register data have not been validated, as done in EUROCAT, consequently there may be a small portion of children reported with clubfoot (ICD-10-DK codes DQ660 and DQ663B) that were not diagnosed with congenital clubfoot [4, 22]. This reflects a clinical reality, in that these are the children referred to specialized care on the basis of their clinical presentation and is not expected to materially alter the findings of the study. Furthermore, an assessment of clubfoot registration in the Danish National Patient Register compared to the number of isolated clubfoot cases reported to the Register of Inborn Malformations (operative from 1983-1994) showed that less than 1% were not registered in the Danish National Patient Register [16]. Additional limitations of the study were the inability to examine laterality, familial occurrence, or other potential risk factors as these data were not available through the Danish Biobank Register. Lastly, although twins occurred at a rate of 3.8% among live births during the study period, we could not distinguish between singleton and multiple births in our cohort.

## Conclusion

In conclusion, the use of the publicly available Danish Biobank Register provides complete nationwide coverage with specific focus on individuals who were sampled for neonatal screening during their first few days of life. These data are captured within a real-world clinical care setting without risk of ascertainment bias and provide access to diagnostic data for virtually all infants born in Denmark. Furthermore, data stored in the Danish Biobank Register indicates the presence of dried blood spot samples for these infants, allowing for future molecular characterization of case children. Additionally, Danish personal identification numbers enable the linking of individual level data across the many, extensive national registers.

The annual prevalence of clubfoot was relatively stable over three decades and remained stable following the establishment of four clubfoot specialized centers in 2010. The characterization of this nationwide clubfoot cohort provides a resource for future etiopathogenic investigations and life course surveillance of clinical outcomes.

## Data Availability

All data produced in the present study are available upon reasonable request to the authors

## List of abbreviations

aPRR: adjusted prevalence rate ratios
CI: 95% confidence intervals
EUROCAT: the European network of congenital anomalies registers
ICD-10-DK: International Statistical Classification of Diseases and Related Health Problems, 10th revision, Danish version
MCA: multiple congenital anomalies
PRR: prevalence rate ratios

## Declarations

### Ethics approval and consent to participate

Per Danish law and regulations, no formal approval or review of ethics were required for our study as individual patient data were not included. Data were retrieved from publicly available sources, which adhere to General Data Protection Regulations and limits reporting of data to groups greater than five [10].

## Availability of data and materials

All data used in this study are publicly available from the sources referenced. No new data were created or analyzed.

## Author contributions

PLH and MC conceived of the study, PAR, ULT, and KH contributed to the design of the study, PLH contributed substantially to the acquisition of data, performed the statistical analysis, and drafted the article, ULT, KMC, KH, PAR, and MC critically reviewed the article draft, and all authors contributed to the interpretation of results. All authors read and approved the final manuscript.

## Acknowledgments

The authors are grateful to Dr Marie Bækvad-Hansen from the Department for Congenital Disorders, Statens Serum Institut, for help in clarifying practices at the Danish Neonatal Screening Program and Dr Bartlomiej Wilkowski and Dr Steven Chong at the Department of Supply / Digital Infrastructure, Statens Serum Institut for guidance and assistance in using the Danish Biobank Register online interface.

