## Supplemental Materials for "Trends in congenital clubfoot prevalence and co-occurring anomalies during 1994-2021 in Denmark: A nationwide register-based study of 1,315,282 live born infants"

**Supplementary Table 1:** List of all major congenital anomalies and syndromes (defined in accordance with EUROCATs classification of malformations <https://eu-rd-platform.jrc.ec.europa.eu/system/files/public/eurocat/Guide_1.5_Chapter_3.3.pdf>) which were assessed within the 1994-2021 Danish clubfoot cohort. The Danish [Classification of diseases and health-related conditions] ICD-10 codes (ICD-10-DK) (listed here: <https://medinfo.dk/sks/brows.php?s_nod=6199>) were used to define the major congenital anomalies and syndromes investigated and are listed below. Conditions were assessed hierarchically in the order listed.

| **Malformation Group** | **Description** | **Included codes (incl subgroups)** | **Excluded minor malformations** |
| --- | --- | --- | --- |
| Clubfoot cases | Talipes equinovarus & Clubfood, unspecified | DQ660, DQ663B |  |
| EXCLUDE | Anencephaly and similar malformations | DQ00 |  |
|  | Encephalocele | DQ01 |  |
|  | Spina bifida | DQ05 |  |
|  | Bilateral renal agenesis | DQ601 |  |
|  | Potter sequence | DQ606 |  |
|  | Arthrogryposis multiplex congenita | DQ743 |  |
| Group C - CHROMOSOMAL ANOMALIES | Down syndrome (trisomy 21) | DQ90 |  |
|  | Edwards syndrome (trisomy 18) & Patau syndrome (trisomy 13) | DQ91 |  |
|  | Triploidy and polyploidy | DQ92 |  |
|  | Monosomies and deletions from the autosomes, not elsewhere classified | DQ93 | DQ936 |
|  | Turner syndrome | DQ96 |  |
|  | Other sex chromosome abnormalities, female phenotype, not elsewhere classified | DQ97 |  |
|  | Other sex chromosome abnormalities, male phenotype, not elsewhere classified | DQ98 |  |
|  | Other chromosome abnormalities, not elsewhere classified | DQ99 |  |
| Group G - GENETIC SYNDROMES, SKELETAL DYSPLASIAS, AND MONOGENIC DISORDERS | Di George syndrome | DD821 |  |
|  | Alagille syndrome | DQ447B |  |
|  | Polycystic kidney disease | DQ611, DQ612, DQ613 |  |
|  | Meckel-Gruber syndrome | DQ619A |  |
|  | Cleidocranial dysostosis | DQ740B |  |
|  | Craniofacial dysostosis | DQ751 |  |
|  | Mandibulofacial dysostosis | DQ754 |  |
|  | Osteochondrodysplasia with defects of growth of tubular bones and spine | DQ77 |  |
|  | Other osteochondrodysplasias | DQ780-DQ788 |  |
|  | Ehlers-Danlos syndrome | DQ796 |  |
|  | Congenital malformations of the skin | DQ800-DQ824 |  |
|  | Neurofibromatosis (nonmalignant) | DQ850 |  |
|  | Tuberous sclerosis | DQ851 |  |
|  | Struge-Weber(-Dimitri) syndrome | DQ858C |  |
|  | Other specified congenital malformation syndromes affecting multiple systems | DQ87 | DQ870D, DQ870F, DQ870G, DQ870I, DQ872E, DQ872G |
|  | Kartagener's syndrome | DQ893F |  |
|  | Deletions seen only at prometaphase | DQ936 |  |
| Group T - TERATOGENIC SYNDROMES | Congenital malformation syndromes due to known exogenous causes, not elsewhere classified | DQ86 |  |
|  | Congenital rubella syndrome | DP350 |  |
|  | Congenital cytomegalovirus infection | DP351 |  |
|  | Other congenital viral diseases | DP358 |  |
|  | Congenital toxoplasmosis | DP371 |  |
| Group M.1 - CARDIAC MALFORMATIONS | Congenital malformations of cardiac chambers and connections | DQ20 |  |
|  | Ventricular septal defect | DQ210 |  |
|  | Atrial septal defect | DQ211 | DQ211C |
|  | Atrioventricular septal defect | DQ212 |  |
|  | Tetralogy of Fallot | DQ213 |  |
|  | Aortopulmonary septal defect | DQ214 |  |
|  | Other congenital malformations of cardiac septa | DQ218 |  |
|  | Congenital malformation of cardiac septum, unspecified | DQ219 |  |
|  | Congenital malformations of pulmonary and tricuspid valves | DQ22 |  |
|  | Congenital malformations of aortic and mitral valves | DQ23 |  |
|  | Other congenital malformations of heart | DQ24 | DQ246 |
|  | Congenital malformations of great arteries | DQ25 | DQ254E |
|  | Congenital malformations of great veins | DQ26 | DQ261 |
| Group M.2 - NERVOUS SYSTEM DEFECTS | Microcephaly | DQ02 |  |
|  | Congenital hydrocephalus | DQ03 |  |
|  | Other congenital malformations of brain | DQ04 |  |
|  | Other congenital malformations of spinal cord | DQ06 |  |
|  | Other congenital malformations of nervous system | DQ07 | DQ078D, DQ078G |
|  | Cyclopia | DQ870D |  |
| Group M.3 - URINARY SYSTEM DEFECTS | Renal agenesis and other reduction defects of kidney | DQ60 |  |
|  | Cystic kidney disease | DQ61 | DQ610 |
|  | Congenital obstructive defects of renal pelvis and congenital malformations of ureter | DQ62 | DQ627 |
|  | Other congenital malformations of kidney | DQ63 | DQ633 |
|  | Other congenital malformations of urinary system | DQ64 |  |
|  | Prune belly syndrome | DQ794 |  |
| Group M.4 - ORO-FACIAL CLEFTS | Cleft palate | DQ35 | DQ357 |
|  | Cleft lip | DQ36 |  |
|  | Cleft palate with cleft lip | DQ37 |  |
| Group M.5 - LIMB ABNORMALITIES | Other congenital musculoskeletal deformities | DQ68 | DQ680, DQ682A, DQ683 -DQ685 |
|  | Polydactyly | DQ69 |  |
|  | Syndactyly | DQ70 |  |
|  | Reduction defects of upper limb | DQ71 |  |
|  | Reduction defects of lower limb | DQ72 |  |
|  | Reduction defects of unspecified limb | DQ73 |  |
|  | Other congenital malformations of limb(s) | DQ74 | DQ740G |
| Group M.6 - GENITAL ANOMALIES | Congenital malformations of ovaries, fallopian tubes and broad ligaments | DQ50 | DQ501, DQ502, DQ505 |
|  | Congenital malformations of uterus and cervix | DQ51 |  |
|  | Other congenital malformations of female genitalia | DQ52 | DQ523, DQ525, DQ527 |
|  | Hypospadias | DQ54 | DQ544 |
|  | Other congenital malformations of male genital organs | DQ55 | DQ552B, DQ552F |
|  | Indeterminate sex and pseudohermaphroditism | DQ56 |  |
| Group M.7 - GASTRO-INTESTINAL ANOMALIES | Other congenital malformations of tongue, mouth and pharynx | DQ38 | DQ381, DQ382, DQ385B |
|  | Congenital malformations of oesophagus | DQ39 |  |
|  | Other congenital malformations of upper alimentary tract | DQ40 | DQ400, DQ401 |
|  | Congenital absence, atresia and stenosis of small intestine | DQ41 |  |
|  | Congenital absence, atresia and stenosis of large intestine | DQ42 |  |
|  | Other congenital malformations of intestine | DQ43 | DQ430 |
|  | Congenital malformations of gallbladder, bile ducts and liver | DQ44 | DQ444 |
|  | Other congenital malformations of digestive system | DQ45 | DQ458B |
|  | Congenital diaphragmatic hernia | DQ790 |  |
| Group M.8 - EYE ABNORMALITIES | Congenital malformations of eyelid, lacrimal apparatus and orbit | DQ10 | DQ101, DQ102, DQ103, DQ105 |
|  | Anophthalmos, microphthalmos and macrophthalmos | DQ11 |  |
|  | Congenital lens malformations | DQ12 |  |
|  | Congenital malformations of anterior segment of eye | DQ13 | DQ135 |
|  | Congenital malformations of posterior segment of eye | DQ14 |  |
|  | Other congenital malformations of eye | DQ15 |  |
| Group M.9 - EAR, FACE, AND NECK ABNORMALITIES | Congenital malformations of ear causing impairment of hearing | DQ16 |  |
|  | Other congenital malformations of ear | DQ17 | DQ170-DQ175, DQ179 |
|  | Other congenital malformations of face and neck | DQ18 | DQ180-DQ182, DQ184-DQ187, DQ189 |
| Group M.10 - ABDOMINAL WALL DEFECTS | Omphalocele | DQ792 |  |
|  | Gastroschisis | DQ793 |  |
|  | Other congenital malformations of abdominal wall | DQ795 |  |
| Group M.11 - RESPIRATORY ANOMALIES | Choanal atresia | DQ300 | DQ309 |
|  | Congenital malformations of trachea and bronchus | DQ32 | DQ320; DQ322 |
|  | Congenital malformations of lung | DQ33 | DQ331 |
|  | Other congenital malformations of respiratory system | DQ34 |  |
| Group M.12 - OTHER ANOMALIES | cystic hygroma | DD181A |  |
|  | Isomerism of atrial appendages | DQ206 |  |
|  | Dextrocardia | DQ240 |  |
|  | Other congenital malformations of skull and face bones | DQ750 |  |
|  | Polands syndrom | DQ798S |  |
|  | Moebius syndrome | DQ870G |  |
|  | Pierre Robin sequence | DQ870I |  |
|  | Sirenomelia syndrome | DQ872E |  |
|  | VACTERL association | DQ872G |  |
|  | Congenital malformations of spleen | DQ890 |  |
|  | Situs inversus | DQ893 |  |
|  | Conjoined twins | DQ894 |  |
|  | Caudal dysplasia sequence | DQ898T1 |  |

**Supplementary Figure 1:** A comparison of the prevalence of primary clubfoot in children born in Denmark with respect to the number of their parents born in Denmark. The number of primary clubfoot cases are indicated above the bars, error bars indicate the 95% confidence intervals.

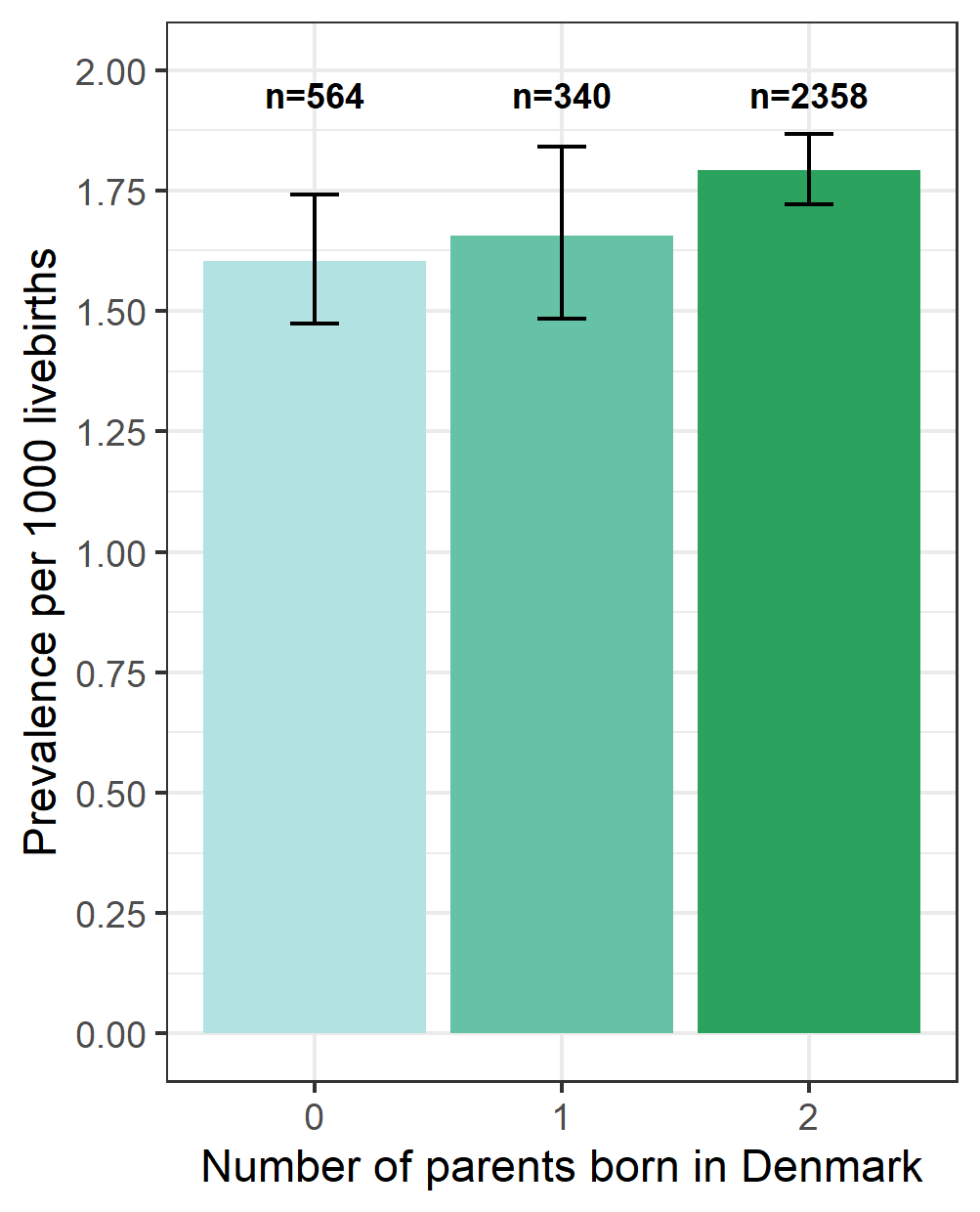

**Supplementary Figure 2:** Trend in the percentage of women who report using tobacco products during pregnancy. Policies and regulations implemented to reduce smoking in Denmark are displayed as labels at the year of their adoption, amendment, or repeal.

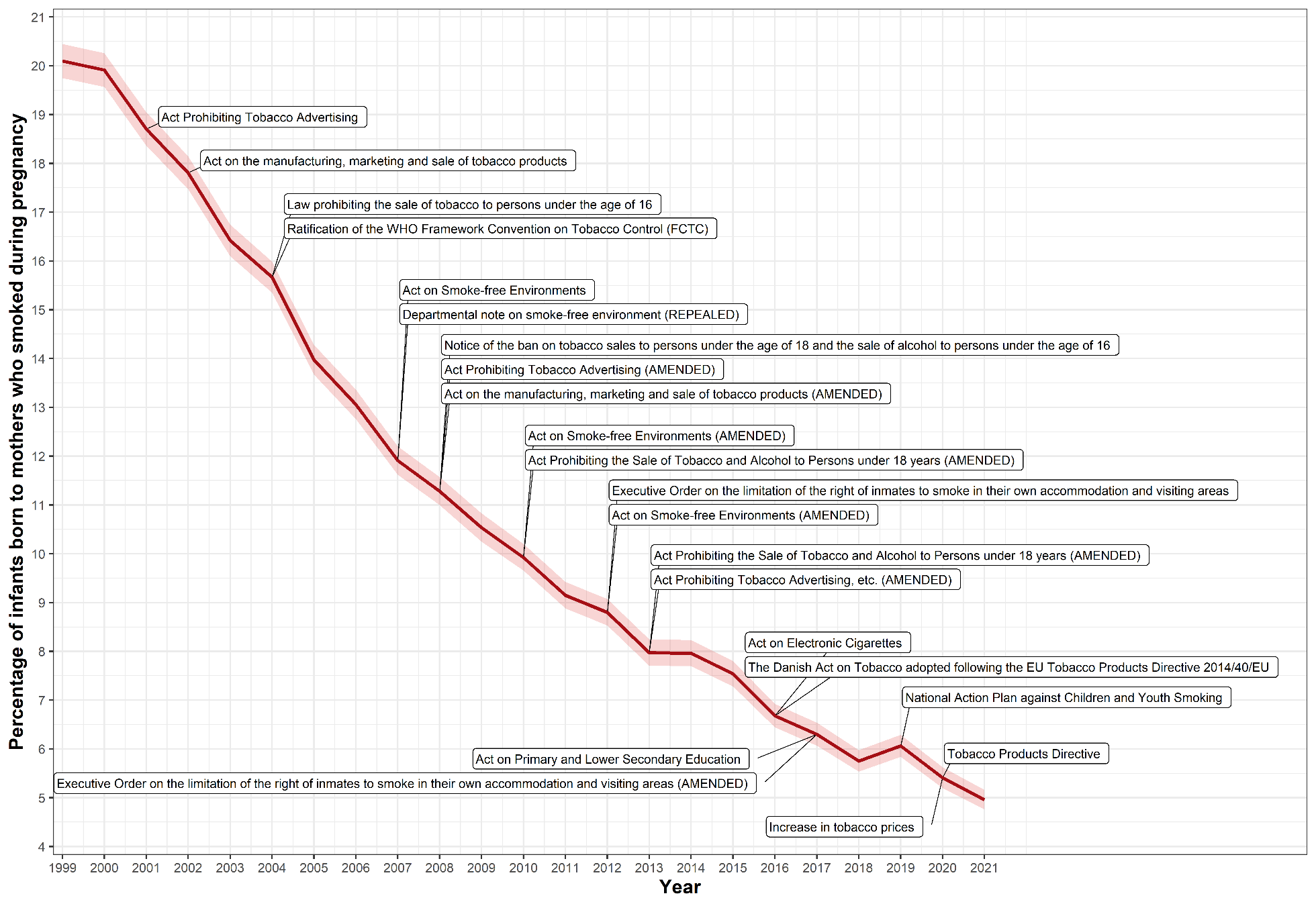
